# Qualitative feasibility study of the mobile app Destroke for clinical stroke monitoring based on the NIH Stroke Scale

**DOI:** 10.1101/2023.04.10.23288374

**Authors:** Evan K Noch, Dan Pham, Tomoko Kitago, Marissa Wuennemann, Susan Wortman-Jutt, M. Cristina Falo

## Abstract

**Background:** Stroke is a leading cause of severe disability in the United States, but there is no effective method for patients to accurately detect the signs of stroke at home. We developed a mobile app, Destroke, that allows remote performance of a modified NIH stroke scale (NIHSS) by patients.

**Aims:** To assess the feasibility of a mobile app for stroke monitoring and education by patients with a history of stroke

**Materials and Methods:** We enrolled 25 patients with a history of stroke in a prospective open-label study to evaluate the feasibility of the Destroke app in patients with stroke. Nineteen patients completed all study assessments, with a median time from stroke onset to enrollment of 5.6 years (range 0.1-12 years). We designed a modified NIHSS that assessed 12 out of 16 tasks on the NIHSS. Patients completed this test eight times over a 28-day period. We conducted pre-study surveys that assessed demographic information, stroke and cardiovascular history, baseline NIHSS, and experience using mobile technologies, and mid- and post-study surveys that assessed patient satisfaction on app usage and confidence in stroke detection.

**Results:** Ten men and nine women participated in this study (median age of 64 (33-76)), representing ten US states and Washington D.C. Median baseline NIHSS was 0 (0-4). 15 patients reported using health apps. On a 5-point Likert scale, patients rated the app as 4.2 on being able to understand and use the app and 4.3 on using the app when instructed by their doctor. For eight patients with poor confidence in detecting the signs of a stroke before the study, six showed higher confidence after the study.

**Conclusions:** The use of an at-home stroke monitoring app is feasible by patients with a history of stroke and improves confidence in detecting the signs of stroke.

## Introduction

Stroke is the leading cause of severe disability and mortality in the US, affecting 750,000 new patients each year^1, 2^. Time to treatment (TTT) is a critical determinant of stroke outcome, with the greatest potential for burden reduction when patients are seen within six hours of symptom onset^3^. However, patients, their loved ones, and caregivers often fail to detect the clinical signs of stroke accurately and efficiently at home, resulting in increased TTT and worse clinical outcomes.

National stroke awareness campaigns such as BEFAST for Stroke have encouraged the public to recognize and react quickly to facial droop, slurred speech, and other stroke signs^4^. However, even when applied diligently, prehospital stroke scales rely on subjective recognition of these signs, and they fail to provide a comprehensive analysis of the full spectrum of stroke presentations. Studies have also shown that these campaigns only reduce the average TTT for patients while the campaigns are active^5, 6^; the improvement in TTT is lost as soon as patients’ awareness wanes.

Destroke is a patient-centered mobile app that uses automated speech, motion, and facial recognition technologies to perform a modified NIH stroke scale (NIHSS). This app is designed to improve stroke education by showing patients the components of the NIHSS and encouraging regular engagement with the app to reinforce awareness of stroke signs. Though mobile technologies such as Destroke may not be feasible for patients with severe cognitive or physical limitations, this app is intended to be used by patients with minor strokes, who traditionally wait longer for treatment than those patients presenting with more severe stroke symptoms^7^. Because the app uses automated analysis of stroke signs, it is designed to minimize the subjectivity associated with self-interpretation of acute neurological signs.

To test the feasibility of stroke patients using the Destroke app at home, we conducted a prospective, open-label study in patients with a history of stroke who used this app over a one-month period at home. Patients performed the Destroke modified NIH-based stroke test twice per week for four weeks, for a total of eight tests. They also answered a 36-question survey at the beginning of the study to assess their stroke history and usage of mobile technologies and a satisfaction survey composed of 36 questions at the midpoint and endpoint of the study.

## Methods

### Standard Protocol Approvals, Registrations, and Patient Consents

We conducted a prospective, open-label, IRB-approved single-center qualitative study of patients with stroke (19-04020233). This study was approved by the Biomedical Research Alliance of New York Institutional Research Board. All patients provided written consent to participate in the study.

### Study Procedures

Recruitment for the study included/was done via ResearchMatch, a national health volunteer registry that was created by several academic institutions and supported by the U.S. National Institutes of Health as part of the Clinical Translational Science Award (CTSA) program. ResearchMatch has a large population of volunteers who have consented to be contacted by researchers about health studies for which they may be eligible. Review and approval for this study and all procedures was obtained from Biomedical Research Alliance of New York. We also recruited patients through social media, a local research registry at the Burke Neurological Institute, and flyers sent to physicians and therapists in the community.

Male or female patients ≥ 18 years of age with evidence of a prior stroke (ischemic stroke, intracerebral hemorrhage, or subarachnoid hemorrhage based on magnetic resonance imaging (MRI) brain scan or computed tomography (CT) scan were included. Patients unable to complete an NIHSS examination through use of the app, including those with impaired alertness, severely impaired comprehension or attention, impaired visual acuity (even with correction), or with bilateral arm plegia, were excluded.

After recruitment, patients participated in a one-hour Zoom session, where the following procedures were performed: informed consent, collection of demographic information, baseline NIHSS, pre-study survey, and baseline test on the Destroke app. Patients were then given instructions to perform the Destroke test three additional times before Day 14, at which point they were instructed to take the mid-study survey. They were instructed to take the Destroke test four additional times until Day 28, when they were instructed to take the post-study survey. In total, patients were asked to perform the Destroke scale eight times over a 28-day period.

From the pre-study survey, we compiled data on age, gender, race, ethnicity, date of prior stroke(s), time to seek medical assistance from stroke, living situation, smartphone ownership and usage, and mobile app and health app usage. We also compiled data on confidence level in detecting stroke on a 5-point Likert scale. The mid- and post-study surveys were identical. In these studies, we compiled data on the following features based on a 5-point Likert scale: time to complete the Destroke scale, instances of app malfunctions, ease of understanding and use, usability of individual Destroke components, engaging nature of the app, likelihood of using the app at various time intervals or when instructed by a doctor, the extent to which the app made patients feel in control of stroke recovery, reassured about stroke symptoms, confident in stroke detection, and reassured if loved ones used the app. We also asked patients how satisfied they were in the app’s ability to help them understand their stroke symptoms.

The Destroke app is a mobile app that incorporates 12 out of the 16 tasks on the NIHSS into the Destroke scale by asking the following questions:

- How old are you?
- What month is it?
- Move your eyes to the left and right.
- Smile and show your teeth.
- Raise your right arm.
- Raise your left arm.
- Raise your right leg.
- Raise your left leg.
- Can you feel this vibration?
- Name these objects.
- Read these sentences.

The four tests that are not assessed are level of consciousness, visual fields, neglect, and ataxia. Level of consciousness is not directly assessed because users must be conscious to operate the app. Visual fields are difficult to assess using a single mobile phone because of the small size of the screen and difficulty in patients seeing the screen adequately when the phone is in their peripheral visual field. Likewise, neglect cannot be adequately assessed with a single mobile phone because of the need to test double simultaneous simulation. Algorithms for ataxia have not been adequately developed to assess abnormalities of coordinated movement when holding the mobile phone. Though dysarthria is not directly tested in the app, word clarity is detected based on algorithms in the app.

All modalities of the Destroke app, except for arm and leg movement, were assessed by a member of the research team using visual and audio recordings from the app. For vibration assessment, the patient holds the phone in each hand, and the phone vibrates. Movement assessment was performed automatically by the app based on accelerometry data. The test and results screens of the Destroke app are shown in Figure 1, and instructional videos for raising arms and raising legs are shown in Supplementary Videos 1 and 2, respectively. For app crashes, patients were instructed to close the app and re-open it, or to contact the study investigators for assistance.

**Figure 1.**
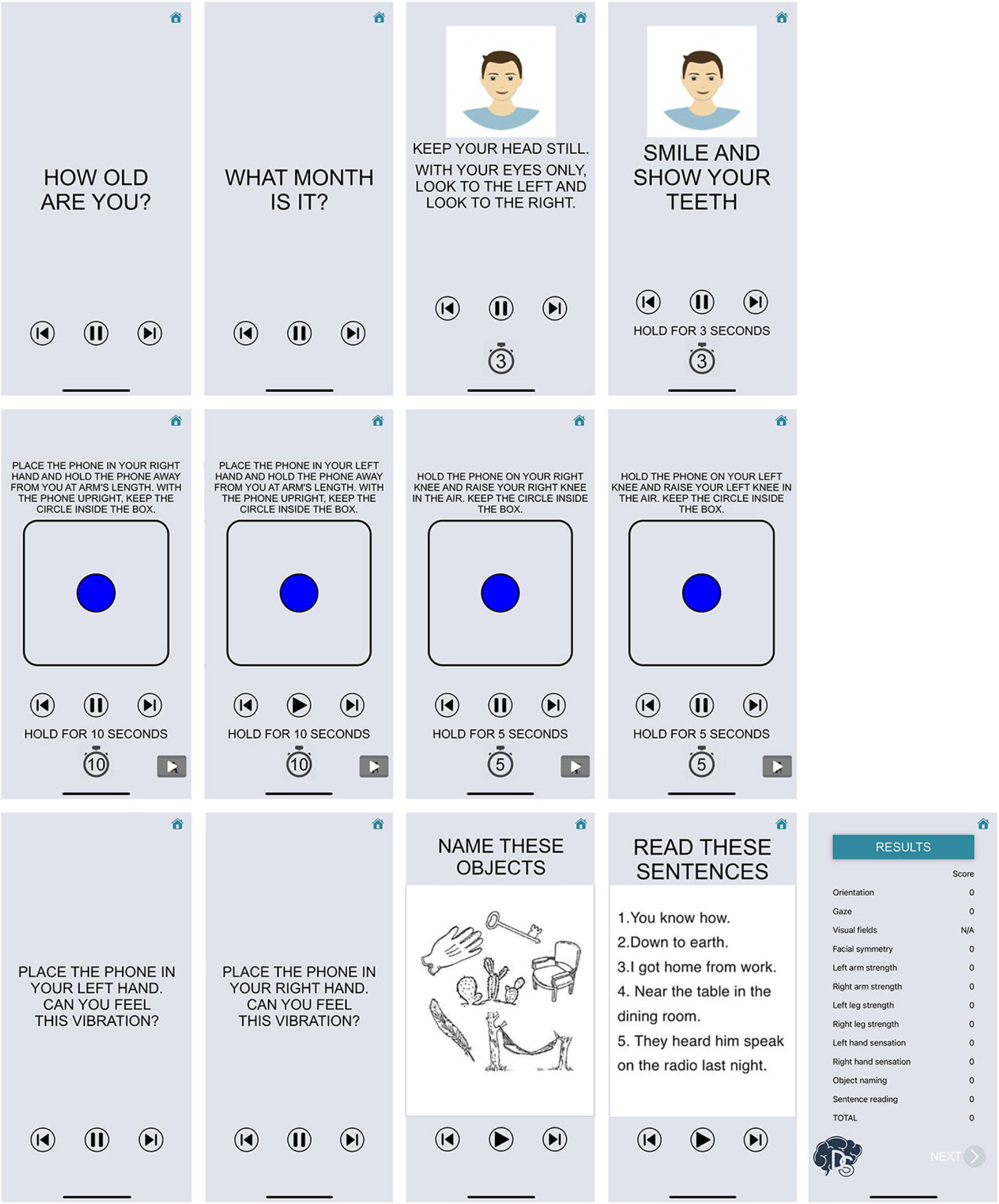
Destroke app test and results screens.

### Statistical analyses

Statistical analyses, including linear regression, were conducted using GraphPad Prism version 8.0 (La Jolla, CA, USA).

## Results

We enrolled 25 patients in this study, and 19 patients completed all study procedures (Figure 2). Three patients were lost to follow-up, two patients voluntarily withdrew from the study, and one patient’s iPhone could not use its voice command function, Siri. There were ten men and nine women who completed the study (Table 1). The median age was 64 (33-76). 16 patients identified as white, two as African American, and one as other. One patient identified as Hispanic. Patients came from ten states (Arizona, Connecticut, Iowa, New York, Ohio, Pennsylvania, Tennessee, South Carolina, Washington, Wisconsin) and Washington DC. The mean time from stroke to enrollment was 5.6 years (0.1-12). The median NIHSS at baseline was 0 (0-4). Three patients had mild sensory loss, three patients had mild neglect, and two patients had mild hemianopsia. One patient each had mild facial droop, mild aphasia, mild dysarthria, and mild ataxia. One patient had no movement of her right leg. No patients had neurological deficits that precluded their use of their mobile phone or the Destroke app.

**Figure 2.**
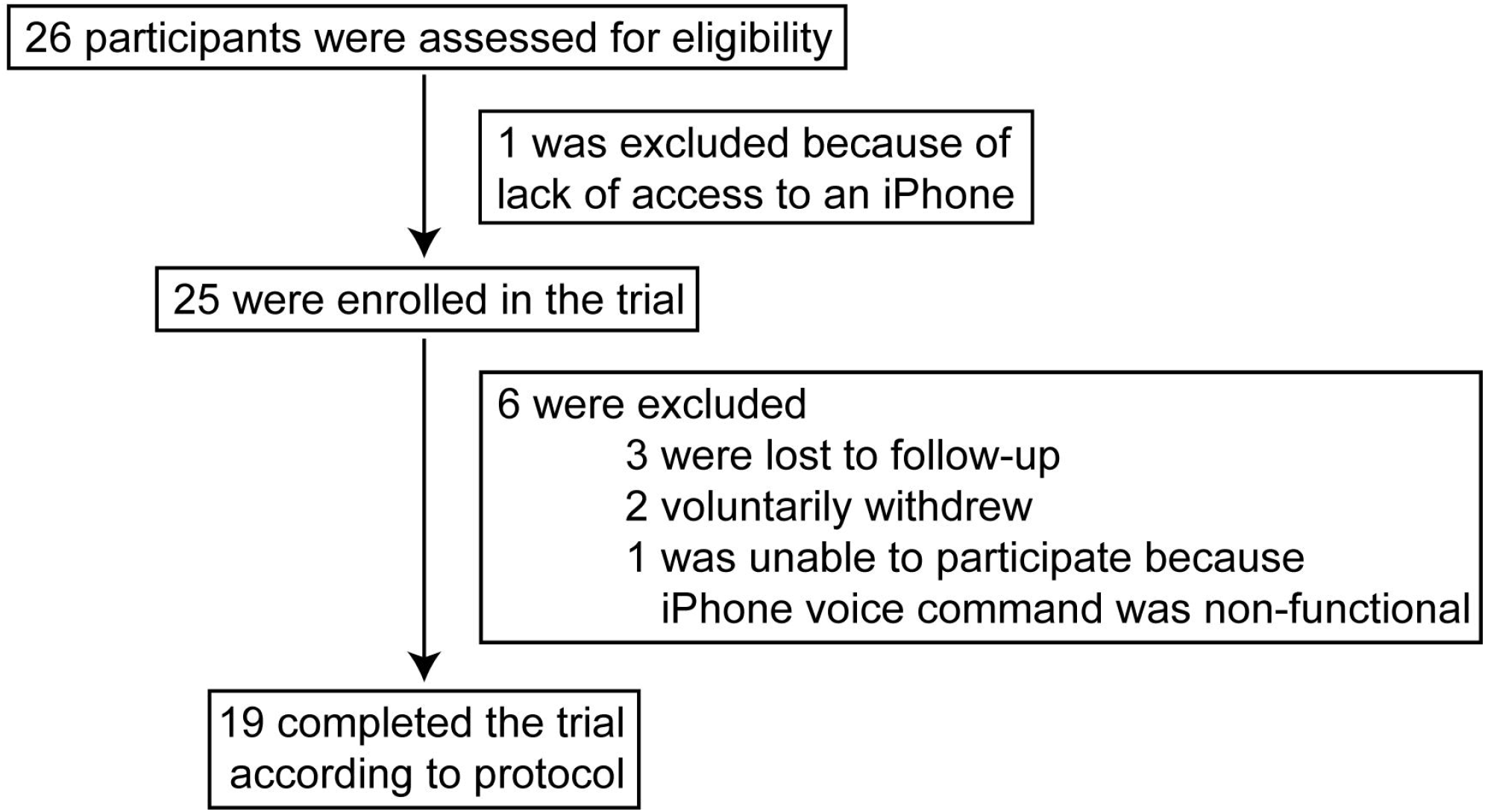
Assessment for eligibility.

**Table 2.**
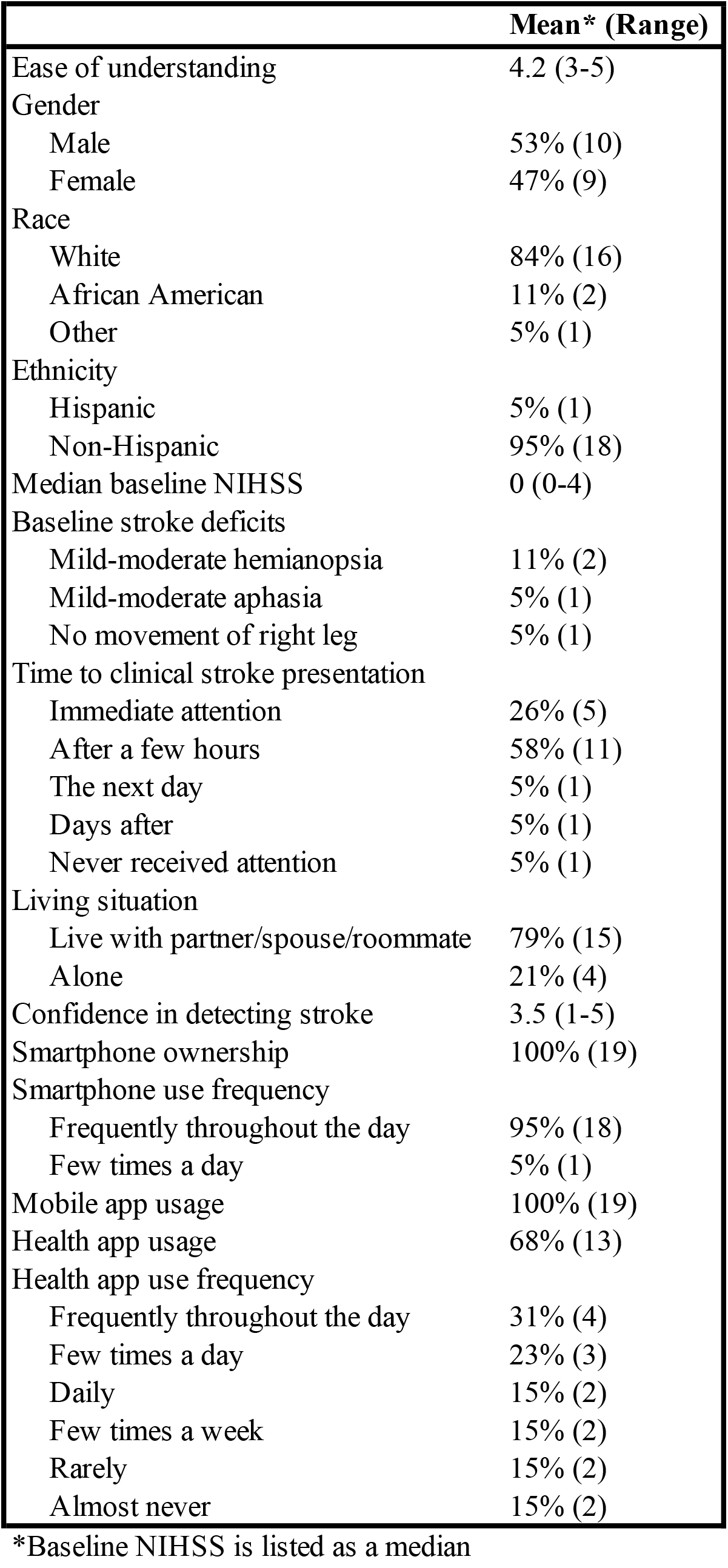
Patient satisfaction scores

| **Table 2** Patient satisfaction scores | **Mean\* (Range)** |
| --- | --- |
| Ease of understanding | 4.2 (3-5) |
| Gender |  |
| Male | 53% (10) |
| Female | 47% (9) |
| Race |  |
| White | 84% (16) |
| African American | 11% (2) |
| Other | 5% (1) |
| Ethnicity |  |
| Hispanic | 5% (1) |
| Non-Hispanic | 95% (18) |
| Median baseline NIHSS | 0 (0-4) |
| Baseline stroke deficits |  |
| Mild-moderate hemianopsia | 11% (2) |
| Mild-moderate aphasia | 5% (1) |
| No movement of right leg | 5% (1) |
| Time to clinical stroke presentation |  |
| Immediate attention | 26% (5) |
| After a few hours | 58% (11) |
| The next day | 5% (1) |
| Days after | 5% (1) |
| Never received attention | 5% (1) |
| Living situation |  |
| Live with partner/spouse/roommate | 79% (15) |
| Alone | 21% (4) |
| Confidence in detecting stroke | 3.5 (1-5) |
| Smartphone ownership | 100% (19) |
| Smartphone use frequency |  |
| Frequently throughout the day | 95% (18) |
| Few times a day | 5% (1) |
| Mobile app usage | 100% (19) |
| Health app usage | 68% (13) |
| Health app use frequency |  |
| Frequently throughout the day | 31% (4) |
| Few times a day | 23% (3) |
| Daily | 15% (2) |
| Few times a week | 15% (2) |
| Rarely | 15% (2) |
| Almost never | 15% (2) |

\*Baseline NIHSS is listed as a median

Regarding their prior stroke, seven patients reported being alone when they had their stroke. Five patients reported receiving immediate medical attention, 11 patients reported receiving attention after a few hours, one patient received attention the next day, one patient received attention days after, and one patient never received attention. Four patients reported living alone, and fifteen patients reported living with a partner, spouse, family, or roommate. Patients reported an average confidence level of 3.5 out of 5 in their ability to detect a stroke. All patients reported owning a smartphone, and all but one patient reported using their smartphone frequently throughout the day. This patient reported using their smartphone a few times a day. All patients reported using mobile apps, and all but one patient reported hearing about apps to track their health. 15 patients used health apps, with these apps being Apple Health, FitBit, Calm, MyFitnessPal, Migraine Buddy, blood sugar tracker, blood pressure tracker, and period tracker. Of those 15 patients who reported using health apps, four reported using them frequently throughout the day, three used them a few times a day, two used them daily, two used them a few times a week, two used them rarely, and two used them almost never. Patients reported an average score of 4.2 out of 5 that they would use an app to detect a stroke, which increased to 4.7 if this app was recommended by their doctor.

When analyzing results from the post-study survey, six patients said that the app test took less than 3 minutes to complete, twelve patients said that it took 3-5 minutes to complete, and one patient said that it took 6-7 minutes to complete. Nine patients reported that the app crashed at least once. When the app crashed, five patients restarted the app, two patients communicated with the study coordinator, and two patients did not complete the test.

Patients rated the app as 4.2 out of 5 for being able to understand and use the app (Figure 3). When patients were asked how strongly they felt about using the app in certain situations, they rated an average of 2.9 out of 5 for using it daily, 3.8 for using it weekly, 3.8 when suspected of having a stroke, and 4.4 when instructed by their doctor (Fig. 3B). Patients rated the app neutrally (3 out of 5) when asked if the app made them feel in control of their stroke recovery (Fig. 3C).

**Figure 3.**
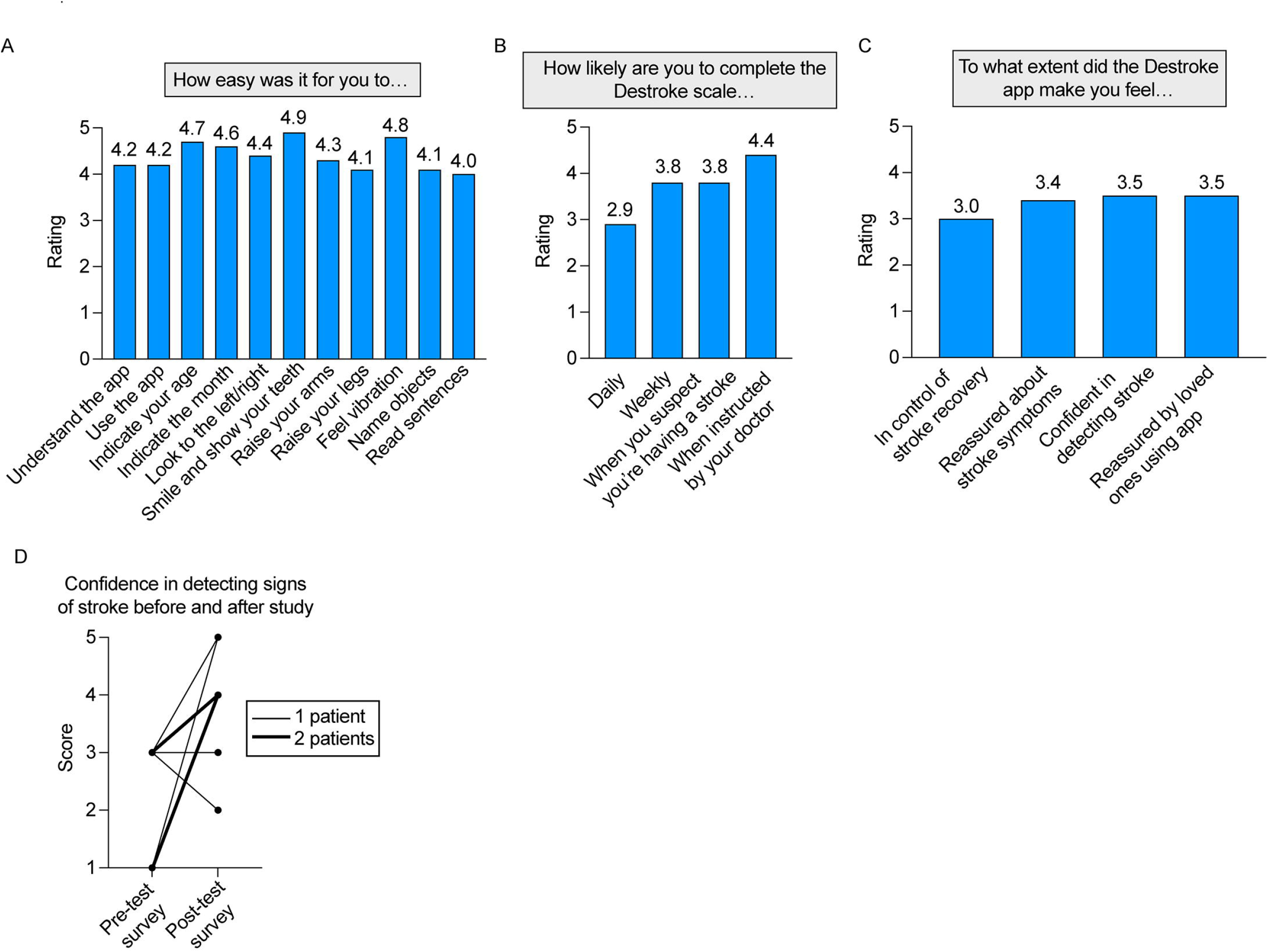
Patient satisfaction data using the Destroke app. (A) Patients indicated their ease of understanding and use of the app and each of its screens. (B) Patients indicated their likelihood of completing the Destroke scale at the indicated intervals. (C) Patients indicated their feelings about the Destroke app. (D) Pre- and post-study confidence in detecting a stroke is shown for patients who rated their confidence at a level of 1-3 prior to the study. (E) Correlation between patients’ confidence in stroke detection and satisfaction in the app helping them to understand their stroke symptoms.

They rated the app 3.4 out of 5 when asked how strongly the app made them feel reassured about their stroke and 3.5 when asked if they felt reassured by loved ones using the app. All but 4 patients felt equally or more confident in their ability to detect the signs of stroke after using the Destroke app as compared to their survey at enrollment. For the eight patients who rated their confidence level in detecting the signs of a stroke as 1-3 before using the Destroke app, six of them showed higher confidence after the study (Fig. 3D). All patients who rated their confidence as 1 before the study rated their confidence level as 4 or 5 after the study. We also found that there was a significant correlation between patients’ confidence in stroke detection and satisfaction in the app helping them to understand their stroke symptoms (R2 = 0.8, p < 0.0001, Figure 3E).

All patients performed between five and nine tests using the app. However, not all tests were complete because of technical difficulties with the app as it was undergoing maintenance or because patients did not permit recording of audio and/or video on their iPhones. We obtained at least one complete Destroke scale from all but two patients, with an average of four out of eight complete scales performed per patient (range 0-7). Baseline and app-scored scales are shown in Figure 4A and B. Baseline NIHSS was adjusted to include only those elements assessed by the app. Therefore, visual fields, ataxia, and neglect scores were removed. In addition, because dysarthria is not directly assessed by the current app, dysarthria scores were removed as well.

**Figure 4.**
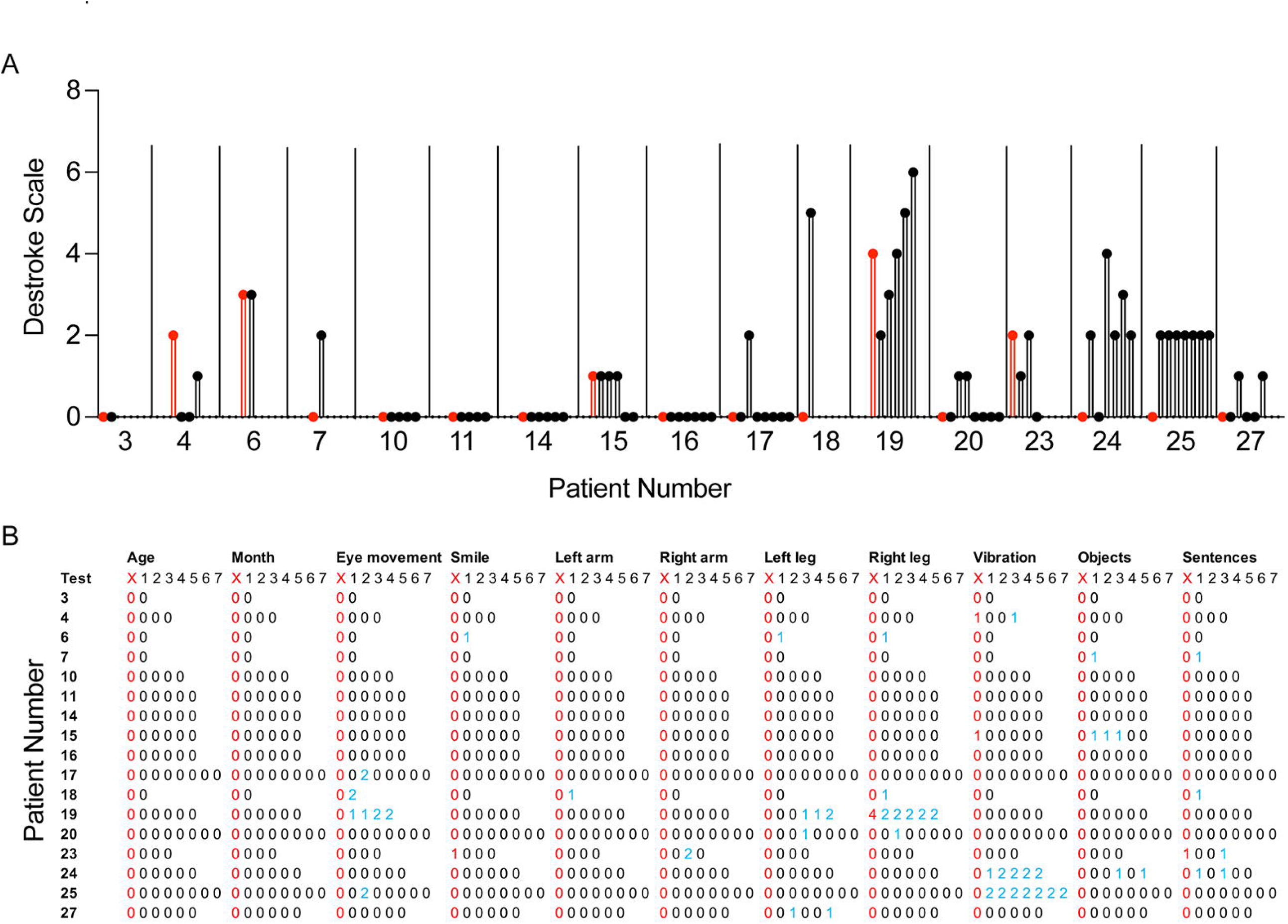
Baseline and app-assessed patient stroke scores. (A) Baseline stroke scores as assessed during a remote video call are shown in red for each patient. Each complete app-assessed stroke score is displayed for each patient. (B) Each stroke score modality is displayed, and scores for each modality on each app-assessed stroke score are shown. The baseline remote video stroke score is shown in red. Non-zero scores on the app are shown in blue.

Patients with aphasia on their baseline NIHSS were counted as having abnormalities on sentence reading. For five patients, app scores matched their baseline scores for all tests. For seven patients, app scores matched the baseline score for at least one test. For five patients, the patient’s baseline score did not match any of the app scores. The most common reasons for lack of completion of any modality or an incorrect response was that the patient did not allow audio or video recording in the app, failure to wait until the vocal command was completed before giving an answer, failure to understand the command, dysfunctional iPhone hardware (e.g. vibration), and app crash. In some of these cases, these issues led to mismatch between the baseline NIHSS and the Destroke scale score. In almost all cases, the NIHSS reported for the app was greater than the baseline NIHSS for the patient. The most common modalities that were not completed were smiling, followed by eye movement, object naming, sentence reading, and vibration reporting.

## Discussion

In this prospective study to evaluate the feasibility of Destroke app usage by patients, we found high degrees of user satisfaction with the overall app and with each task in the app. We found a high likelihood of patients using this app if recommended by their doctors. Importantly, we found that confidence in detecting stroke increased in patients after this 28-day study, and this effect was particularly pronounced for patients who had low confidence levels prior to the study. These findings indicate that the Destroke app may improve patients’ confidence in understanding the signs of stroke.

Based on our findings, patients with a prior stroke use mobile technologies and may engage with mobile apps like Destroke to track their neurological recovery after stroke. The average age of patients in this study was 64, which is only slightly younger than the average age of the stroke population^8^. All patients enrolled in this study reported using a smartphone, and all patients used mobile apps, with most patients (fifteen out of nineteen) using health-related apps. Patients in this study were enrolled from 10 US states and Washington D.C., which minimized the effects of regional differences in smartphone usership.

Prior data indicate that patients who have had a stroke cannot accurately report the signs of stroke^4^. This lack of awareness is present in both inpatient and outpatient settings and even immediately after a recent stroke. National stroke awareness campaigns improve stroke detection by patients and their loved ones, but this is only the case while these campaigns are active^5, 6, 9, 10^. We found that only five patients in our study sought immediate attention for their stroke, indicating a general lack of awareness of the signs of stroke. The higher confidence level achieved for most patients in our study may translate to more rapid recognition of these signs and improved TTT. As the remote health monitoring industry grows, mobile patient-centered technologies could be used to improve stroke education and awareness^11, 12^. These technologies would ideally be oriented to patients at risk for stroke, such as those with prior stroke and/or cardiovascular risk factors, because they may be more invested in stroke prevention. Using in-app reminders to perform regular stroke assessments as well as integration with electronic medical record systems and patient portals, these apps could be designed to maintain stroke awareness over the long-term. Such a paradigm would be more personalized than broad advertising campaigns and may better reach users who engage with their smartphones frequently throughout the day, as reported by all patients in this study.

This study was limited by a small sample size and by a short 28-day duration, preventing ascertainment of the stability of confidence levels in stroke detection after chronic app usage and after app disuse. The Destroke app does not currently test visual fields, coordination, dysarthria, and neglect. In particular, the abilities of a single smartphone without additional external sensors are limited in detection of visual fields and sensory neglect. However, some of these features, such as coordination and dysarthria, will be included in future app versions. The app is also limited in usage to those with relatively low NIHSS and without disorders of consciousness or awareness who are still able to operate mobile technologies. Because this study involves the use of a mobile app, there may also be a strong placebo effect in rating confidence higher after the study intervention. Because the study was conducted remotely, we did not directly observe performance of the Destroke scale after the initial demo, preventing complete understanding of app failures in instances of incomplete Destroke scales. The Destroke app is currently available only on the iOS platform, limiting generalizability to Android users, who comprise about 35% of the US population^13^.

As telehealth usage increases to satisfy the healthcare needs of a more diverse, technology-savvy population, remote monitoring of stroke can enhance comfort of stroke patients in evaluating their stroke symptoms and in monitoring their recovery. Next steps in this work will involve testing the use of the Destroke app in a randomized fashion in those with stroke and in patients with higher baseline NIHSS. We will also examine the stability of stroke symptom awareness with both continuous and less frequent app usage. Further integration of mobile stroke apps with electronic medical record systems and telehealth providers will provide seamless evaluation by patients and providers and may better address current deficiencies in pre-hospital stroke detection and treatment.

## Data Availability

Anonymized data not published within this article will be made available by request from any qualified investigator.

## Data Access Statement

The Principal Author takes full responsibility for the data, the analyses and interpretation, and the conduct of the research. They have full access to all of the data, and they have the right to publish any and all data, separate and apart from the guidance of any sponsor.

## Declarations of conflicting interests

E.K.N. is a founder and CEO of Destroke, which is developing mobile technologies for clinical stroke detection.

## Funding

The author(s) discloses receipt of the following financial support for the research, authorship, and/or publication of this article: Destroke, Inc. and Burke Neurological Institute.

## Lead Contact Statement

Further information and requests for data and resources should be directed to and will be fulfilled by the Lead Contact, M. Cristina Falo.

## Figure Legends

**Supplementary Figure 1 Video demonstrating instructions for raising arms**

**Supplementary Figure 2 Video demonstrating instructions for raising legs**

## Notes

### Author Declarations

This study was approved by the Biomedical Research Alliance of New York Institutional Research Board.

